# Association of working shifts, inside and outside of healthcare, with risk of severe COVID-19: An observational study

**DOI:** 10.1101/2020.12.16.20248243

**Authors:** A.V. Rowlands, C. Gillies, Y. Chudasama, M.J. Davies, N. Islam, D.E. Kloecker, C. Lawson, M. Pareek, C. Razieh, F. Zaccardi, T. Yates, K. Khunti

## Abstract

**Background:** Health and key workers are at an increased risk of developing severe COVID-19; it is not known, however, if this risk is exacerbated in those with irregular work patterns. We aimed to investigate the risk of severe COVID-19 in health and shift workers.

**Methods:** We included UK Biobank participants in employment or self-employed at baseline and with linked COVID-19 data to 31^st^ August 2020. Participants were grouped as neither a health worker nor shift worker (reference category), health worker only, shift worker only, or both and associations with severe COVID-19 investigated in logistic regressions.

**Findings:** Of 235,685 participants (81·5% neither health nor shift worker, 1·4% health worker only, 16·9% shift worker only, and 0·3% both), there were 580 (0·25%) cases of severe COVID-19. The risk of severe COVID-19 was higher in health workers (adjusted odds ratio: 2.32 [95% CI: 1·33, 4·05]; shift workers (2·06 [1·72, 2·47]); and in health workers who worked shifts (7·56 [3·86, 14·79]). Being both a health worker and a shift worker had a possible greater impact on the risk severe COVID-19 in South Asian and Black and African Caribbean ethnicities compared to White individuals.

**Interpretation:** Both health and shift work were independently associated with over twice the risk of severe COVID-19; the risk was over seven times higher in health workers who work shifts. Vaccinations, therapeutic and preventative options should take into consideration not only health and key worker status but also shift worker status.

**Funding:** National Institute for Health Research, UK Research and Innovation.

**Research in context:** *Evidence before this study:* The risk of developing severe COVID-19 is greater in occupational groups with higher levels of viral exposure, e.g. health and key workers. We searched PubMed and medRxiv up to December 8, 2020 for papers on shift work patterns, health work and incidence of COVID-19 using the keywords “COVID-19”, “SARS-CoV-2”, “shift work” “health worker”. Recent evidence suggests shift workers are also at increased risk of severe COVID-19 but it is not clear if the risk is exacerbated in those who work shifts in healthcare.

*Added value of this study:* This study uses data from UK Biobank, a prospective cohort of >500,000 adults aged 40-69 years with baseline assessments between March 2006 and July 2010. Participants’ occupation was categorised according to whether or not they were health workers and/or shift workers at baseline. Results showed that being a health worker, or working shifts, were similarly and independently associated with over twice the population level risk of severe COVID-19 independent of age, sex, ethnicity, deprivation and co-morbidities. The risk was seven times higher in health workers with shift working patterns. The impact of health and shift work tended to be higher in males and in minority ethnic groups, who are already at an increased risk of severe COVID-19. In people over the age of retirement, the risk of developing severe COVID-19 associated with baseline health worker status was no longer apparent, suggesting the risk is likely explained by exposure to the virus. However, the elevated risk associated with baseline shift worker status persisted, albeit attenuated.

*Implications of all the available evidence:* Shift workers are at elevated risk of developing severe COVID-19. The persistence of an elevated risk in people who are now over retirement age, but had a shift worker status at baseline, suggests the risk may not be fully explained by increased exposure to the virus. Vaccination, therapeutic and prevention programmes are being prioritised for health care workers. Our data suggests that shift workers should also be prioritised for these preventive measures.

## INTRODUCTION

The severe acute respiratory syndrome coronavirus 2 (SARS-CoV-2), which causes coronavirus disease-2019 (COVID-19), is a global health threat.^1^ It has led to an unprecedented co-ordinated global research effort to develop and evaluate a range of vaccines. To date, preliminary results are in for three candidate vaccines and priority groups for vaccination have been identified. The provisional priority list in the UK focuses on care home residents and their carers, front-line health and social care workers, and older adults.^2^ The high priority for health workers and care workers is due to the established elevated risk in these groups for infection, development of severe infection and spreading infection.^3–7^ This risk is further increased for health workers from ethnic minorities.^6^ There has been less attention on whether the risk is exacerbated in those with irregular work patterns, i.e. shift work, which is common in health and care.

Working shifts is associated with an increased risk for cardiovascular disease^8–11^ which appears to persist following retirement, although attenuated.^11^ Research suggests risk factors for cardiometabolic diseases^12^ are also risk factors for COVID-19.^13–15^ Further, shift work is associated with alterations in the immune system and an increased risk for viral infections.^16^ In this view, it is not surprising that recent evidence suggests that shift work is associated with an elevated risk of severe COVID-19,^5,17^ and that health care workers on night shifts have a higher risk of in-hospital SARS-CoV-2 infection than those on day shifts.^18^ However, it is not known whether working shifts interacts with health worker status or ethnicity, both of which are independently associated with an elevated risk.^6,19,20^

Shift workers are more likely to have disturbed sleep and variable sleep patterns^21^ leading to disruption of the circadian rhythm. This has been hypothesised to increase the risk of COVID-19 in night shift workers,^22^ but is evident even if the shift pattern does not include night work, likely due to sleep disruption in relation to circadian rhythms,^21^ and may persist in the years following cessation of shift work.^23^ Recent data have suggested that sleep disruption and high variability in sleep timing are associated with the risk of testing positive for COVID-19 and development of severe infection.^15^ Exacerbating this, shift work is common in health workers where exposure to infection with SARS-CoV-2 and risk of developing severe COVID-19 is already relatively high.^6,7^ Therefore, we hypothesise both health workers and shift workers will independently be at an increased risk of severe COVID-19, but this risk will be increased in health workers who are also shift workers. Further, we hypothesise that the increased risk will be evident across ethnic groups and for males and females.

## METHODS

This study is reported as per the Strengthening the Reporting of Observational Studies in Epidemiology (STROBE) guidelines (Supplementary material: Checklist S1) and following a pre-specified protocol (Application Number 36371).^24^

### Study population

For this analysis, we used data from UK Biobank (application 36371), a prospective cohort of >500,000 adults aged 40-69 years.^25^ All baseline assessments were conducted between March 2006 and July 2010. UK Biobank data are linked to national SARS-CoV-2 laboratory test data through Public Health England’s Second Generation Surveillance System.^26^ The data were available from 16^th^ March 2020 to 31^st^ August 2020 and included outcome of the test (positive/negative) and specimen origin (hospital inpatient vs other). Analyses were restricted to those who were alive on 16th March 2020 (the first COVID-19 testing sample date) and to English centres as testing data were initially only for those based in England.

#### Exposure

Participants’ occupation was categorised according to whether or not they were health workers and/or shift workers based on the occupation information reported at baseline. Health care workers were classified based on UK Biobank occupational codes 2211001-2216012. Participants who reported that their work involved shift work “sometimes”, “usually” or “always” were classified as shift workers, while participants who reported that their work “never/rarely” involved shift work were classified as non-shift workers. Shift or health worker status was defined as four mutually exclusive categories: neither (reference category), health worker only, shift worker only, or both health and shift worker. Only participants who reported being in paid employment or self-employed at baseline were asked about shift work. Those without data for shift work and health work status were excluded (Supplementary material, Figure S1).

#### Outcome

Severe COVID-19 was defined as a composite of a positive test result for SARS-CoV-2 from a hospital setting in line with guidance for this dataset,^26^ or death related to the disease (i.e. any death with an ICD-10 code of U07.1 or U07.2 as the primary cause of death on the death certificate). Positive tests in an outpatient setting were removed from this analysis as we were unable to determine whether these ultimately resulted in hospitalisation. Results can thus be interpreted as the overall population level risk of being admitted to hospital with or dying from COVID-19 during the linkage period within UK Biobank. This population level method of assessing risk has been commonly reported within COVID-19 risk factor research, enabling comparison to the literature in terms of how the risk factors assessed compare to other commonly reported risk factors.^13–15,27^

#### Co-variates/confounders

Participant characteristics, including body mass index (BMI), sex, ethnicity (White, South Asian, Black and African Caribbean), deprivation (Townsend score, a composite measure of deprivation based on unemployment, non-car ownership, non-home ownership, and household overcrowding; negative values represent less deprivation), cancer (self-reported), co-morbidities (yes/no; one or more medical condition(s): i.e. cardiovascular, respiratory, renal, neurology, musculoskeletal, haematology, gynaecology, immunology, infections), and smoking status (never, previous current) were collected at the baseline assessment. Age on 16th March 2020 was calculated.

#### Statistical Analysis

Logistic regression was used to identify the risk associated with developing severe COVID-19 in shift workers only, health workers only and both health and shift workers, relative to non-health workers with normal working hours. These four categories are mutually exclusive to facilitate interpretation of the independent and effects of shift and health worker status, and whether their combination provides an additive or multiplicative association. Analyses were carried out overall and stratified by ethnicity and sex. Analyses were adjusted for potential confounders (age, sex, BMI, ethnicity, deprivation, cancer, co-morbidities, smoking status) selected based on current knowledge.

Two sensitivity analyses were carried out: 1) Including as co-variate self-reported sleep duration at baseline; 2) Stratified by retirement age (currently 66 years of age in the UK). People below retirement age at the beginning of the pandemic were assumed most likely to still be working and thus at higher exposure to the virus (individuals with an age at time of COVID-19 test equal to or below 65 years). People above retirement age were assumed to be less likely to be working and thus at lower exposure to the virus (individuals with an age at time of COVID-19 above 65 years). Baseline age was measured as an integer in years.

All analyses were carried out in Stata version 16.0 (StataCorp LLC, TX, USA). Statistical significance was set at the alpha level of .05.

## RESULTS

There were 235,685 participants eligible for inclusion in this analysis (i.e. with information on outcome of severe COVID-19, shift or health worker status, and full co-variate profile), of which 580 (0·25%) had severe COVID-19. Mean participant age was 63·8 years (SD 7·1), BMI 27·2 kg·m^-2^ (SD 4·7), 52·2% were female, and 96·1% were White (Table 1); 81·5% (n=193,135) were neither a shift nor health worker, 16·9% (n=38,738) were a shift worker only, 1·4% (n=3,193) a health worker only, and 0·3% (n=620) both.

**Table 1:** Participant characteristics.

| Characteristic | All (n=235,685) | Severe covid-19<br>(hospitalisation or death) |  |
| --- | --- | --- | --- |
|  |  | Yes<br>(n=580) | No<br>(n=235,105) |
| **Age at test (years)** | 63.8 (7.11) | 63.6 (7.70) | 63.8 (7.10) |
| **Body mass index (kg/m<sup>2</sup>)** | 27.2 (4.68) | 28.7 (5.34) | 27.2 (4.68) |
| **Townsend score** | -1.39 (2.96) | -0.47 (3.39) | -1.39 (2.96) |
| **Sex** |  |  |  |
| Female | 123,127 (52.2) | 275 (47.4) | 122,852 (52.3) |
| **Ethnicity** |  |  |  |
| White | 226,436 (96.1) | 518 (89.3) | 225,918 (96.1) |
| South Asian | 4,345 (1.8) | 30 (5.2) | 4,315 (1.8) |
| Black | 4,904 (2.1) | 32 (5.5) | 4,872 (2.1) |
| **Smoking status** |  |  |  |
| Never | 135,710 (57.6) | 300 (51.7) | 135,410 (57.6) |
| Previous | 75,767 (32.2) | 221 (38.1) | 75,546 (32.1) |
| Current | 24,208 (10.3) | 59 (10.1) | 24,149 (10.3) |
| **Past or current cancer** |  |  |  |
| Yes | 13,957 (5.9) | 39 (6.7) | 13,918 (5.9) |
| **Co-morbidities** |  |  |  |
| Yes | 162,290 (68.9) | 438 (75.5) | 161,852 (68.9) |
| **Shift or health worker** |  |  |  |
| Neither | 193,134 (81.5) | 375 (64.7) | 192,759 (82.0) |
| Shift worker | 38,738 (16.9) | 183 (31.6) | 38,555 (16.4) |
| Health worker | 3,193 (1.4) | 13 (2.2) | 3,180 (1.4) |
| Both | 620 (0.3) | 9 (1.6) | 611 (0.3) |

Values reported are mean (SD) or N (%)
Co-morbidities: yes/no; one or more medical condition(s): i.e. cardiovascular, respiratory, renal, neurology, musculoskeletal, haematology, gynaecology, immunology, infection.
Townsend score: a composite measure of deprivation based on unemployment, non-car ownership, non-home ownership, and household overcrowding; negative values represent less deprivation

After adjustment for potential confounders, a significant association was found between shift worker only (adjusted odds ratios (aOR): 2·06 [95% CI: 1·72, 2·47]) or health worker only (2·32 [1·33, 4·05]) status and risk of severe COVID-19 (Figure 1a). The estimated risk was greatest for individuals who were both a shift and health worker (aOR: 7·56 [3·86, 14·79]). A similar pattern was found when the analysis was stratified by sex (Figure 1a), with a higher estimated association for both shift and health worker status in men (aOR: 10·70 [4·92, 23·28]) than women (aOR: 3·58 [0·88, 14·54]). When the analysis was stratified by ethnicity (Figure 1b), there was a tendency for a greater impact of being both a health worker and a shift worker in South Asian and Black and African Caribbean ethnicities when compared to White, but confidence intervals were large.

**Figure 1:**
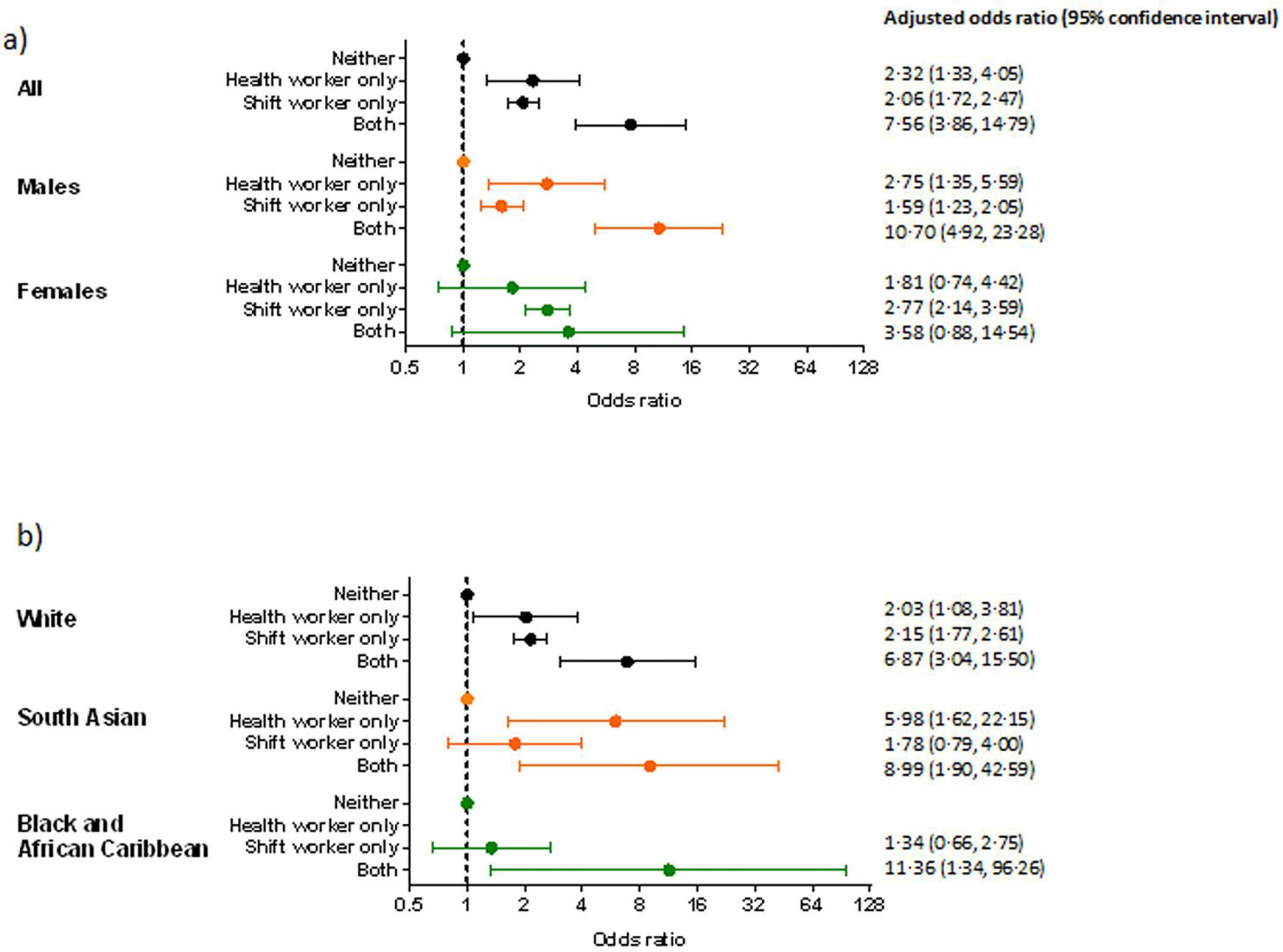
Association between employment status and odds of severe COVID-19 stratified by a) sex and b) ethnicity. Model (a) adjusted for: age, sex (for all participants), Townsend score, BMI, cancer (self-reported, past or current at time of data collection), co-morbidities (yes/no), smoking status (never, previous current) and ethnicity. Model (b) same as model (a) except without ethnicity.

Results of unadjusted models were consistent with the adjusted models and are given in Supplementary Table S1.

### Sensitivity analyses

In the first sensitivity analysis, controlling for sleep duration did not change the results (Supplementary Figure S2).

The second sensitivity analysis, stratified by retirement age, was conducted for the whole sample only, due to small numbers in the sex and ethnicity sub-groups. There were 125,118 eligible individuals below retirement age (54·2% female, 94·8% White) and assumed to be working, of which 312 (0·25%) had severe COVID-19. Of these 80·1% (n=100,170) were neither a shift nor health worker, 18·2% (n=22,819) were a shift worker only, 1·4% (n=1,708) a health worker only, and 0·3% (n=421) both. There was a similar pattern of results, with estimated odds ratios generally larger than when the whole cohort was considered (Figure 2).

**Figure 2:**
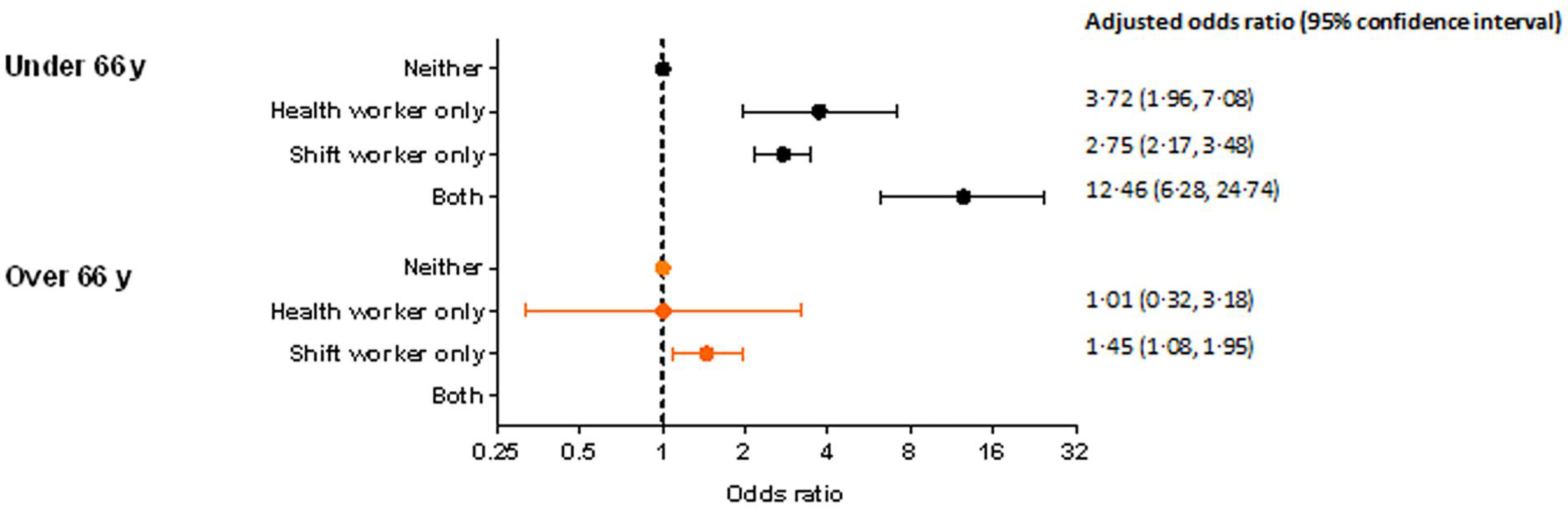
Association between employment status and odds of severe COVID-19 stratified by UK retirement age (66 y) Adjusted for age, sex (for all participants), Townsend score, BMI, cancer (self-reported, past or current at time of data collection), co-morbidities (yes/no), smoking status and ethnicity.

Eligible people above retirement age and assumed to be retired were 110,567 (50·0% female, 97·6% White), of which 268 (0·24%) had severe COVID-19. Of these, 84·1% (n=92,964) had been neither a shift nor health worker, 14·4% (n=15,919) a shift worker only, 1·3% (n=1,485) a health worker only, and none who had been both. The risk associated with prior shift worker status persisted, albeit lower (aOR: 1·45 [1·08, 1·95]); conversely, no risk associated with prior health worker status was evident (Figure 3).

## DISCUSSION

Both being a health worker, or working shifts, were independently associated with over twice the population level risk of severe COVID-19; notably, the risk was more than seven times higher in health workers who work shifts. The impact of health and shift work tends to be higher in males and in minority ethnic groups, who are already at increased risk of severe COVID-19.^19,20^

The risk of severe COVID-19 was stronger when considering only people below retirement age at the beginning of the pandemic, thus more likely to still be working and at increased viral exposure. When considering only people above retirement age at the beginning of the pandemic, the risk associated with health worker status appears to dissipate. This potentially suggests that the elevated risk in the whole population or those under retirement age is indeed explained by increased exposure to the virus. In contrast, an elevated risk associated with prior shift-work status persists, although attenuated. Alongside the higher risk for health workers who work shifts, this suggests that the risk associated with shift work may not be fully explained by an increased viral exposure.

The persistence of an elevated risk associated with shift work following retirement has previously been identified for cardiovascular disease.^11^ Purported mechanisms include disruptions to the behavioural and circadian rhythm,^8^ which can lead to chronic inflammation,^28^ potentially contributing to the increased risk of cardiovascular disease observed in previous shift workers. As COVID-19 is an acute inflammatory disease,^28^ it may exacerbate any existing chronic inflammation. Alongside other risk factors (e.g. lifestyle factors, psychological stress and genetic predisposition), this may be associated with a ‘cytokine storm’^22,28,29^ contributing to the increased risk of severe COVID-19 we observed in shift-workers.

The demand for 24-hour services has extended shift work beyond factories to more traditionally “white collar” occupations, e.g. retail and service,^30^ with approximately 15-25% of workers in Europe employed on shift schedules.^10,16^ Irrespective, shift workers still tend to be more deprived and subject to psychosocial stresses,^9^ which may contribute to increased risk for cardiovascular disease and COVID-19. While we controlled for a range of available co-variates, including age, sex, ethnicity, deprivation, co-morbidities and self-reported sleep (sensitivity analysis), other residual confounders may be present that predispose the shift workers to greater risk. However, Maidstone et al.^17^ recently showed that the incidence of COVID-19 in shift workers was still greater when compared to non-shift workers in the same job. Further, in a previous UK Biobank study, we showed that objectively measured sleep disruption and variability in sleep timing was associated with increased risk of severe COVID-19.^15^ While disturbed sleep is prevalent in shift workers,^21^ the risk was similar when excluding shift workers from the cohort.^15^ This observation would suggest that sleep disturbance and variability in sleep timing, even in the absence of shift work status, is associated with an increased risk. Likewise, irregular sleep timing was associated with metabolic abnormalities in a prospective study on cardiovascular events in ∼2000 participants,^12^ with similar results when shift workers were excluded.

Strengths of this study include the large population with linked COVID-19 data. In addition, the UK Biobank differs from many other datasets currently being analysed to better understand COVID-19, in that it is an extensively phenotyped population, allowing the impact of issues such as shift worker status to be assessed. However, the study also has several important limitations. Characteristics of participants, including health worker and shift work status, were measured between 2006 and 2010. Mutambudzi et al.^5^ and Maidstone et al.^17^ similarly used occupation at UK Biobank baseline to ascertain risk of severe COVID-19. In support of this assumption, Matambudzi et al.^5^ determined a high correlation (r = 0.71, p<0.001) between occupation at baseline and occupation between 2014 and 2019 in a sub-sample of >12,000, participants indicating a high likelihood that participants had continued working in the same profession. Further, in our analyses stratified by retirement age, we assumed that those below retirement age at the date of their COVID-19 test were still working and at relatively high exposure to COVID-19, while those above retirement age were not working and were at lower exposure. It is not possible to confirm this assumption with the available data. Additionally, the definition of severe COVID-19 was a positive test from a hospital inpatient; while this is consistent with the definition proposed by the researchers that developed the linkage method,^26^ actual disease severity cannot be confirmed from the linkage data available. Finally, participants in UK Biobank may not be representative of the wider population and testing in the UK has not been universal, making analyses vulnerable to bias.

In conclusion, both shift and health work were associated with an increased risk of developing severe COVID-19 independent of age, sex, ethnicity, deprivation and co-morbidities. The risk was compounded more than three-fold further in health workers who work shifts, irrespective of sex or ethnicity, compared to neither health nor shift worker. The impact of health and shift work tended to be higher in minority ethnic groups, who are already at increased risk of severe COVID-19. The UK Reach study
https://uk-reach.org/main/) will investigate how, and why, ethnicity affects COVID=19 outcomes in healthcare workers. Notably, the risk associated with health workers was no longer apparent in people over retirement age, suggesting that the risk is likely explained by the exposure to the virus inherent to the occupation. However, in shift workers, an elevated albeit attenuated risk was still evident in people over retirement age, suggesting that the elevated risk associated with shift work may not be fully explained by increased exposure to the virus. This is consistent with previous reports of elevated risk of cardiovascular disease in former shift workers^11^ and further supports that risk factors for cardiovascular and cardiometabolic disease are also risk factors for COVID-19.^13–15^ Vaccination, therapeutic and prevention programmes are being prioritised for health care workers. Our data suggests that shift workers should also be prioritised for these preventive measures.

## Data Availability

: This research has been conducted using the UK Biobank Resource under Application 36371. The data that support the findings of this study are available from UK Biobank project site, subject to registration and application process. Further details can be found at https://www.ukbiobank.ac.uk.

## Contributions

Concept and design: AR, CG, KK, TY

Acquisition, analysis or interpretation of the data: All authors

Statistical analysis and data verification: CG, AR

Drafting of the manuscript: AR, CG

Critical revision of the manuscript for important intellectual content: All authors

## Acknowledgements

Data were analysed using UK Biobank application number 36371.

## Data sharing statement

This research has been conducted using the UK Biobank Resource under Application 36371. The data that support the findings of this study are available from UK Biobank project site, subject to registration and application process. Further details can be found at https://www.ukbiobank.ac.uk.

## Ethics approval

All participants gave written informed consent prior data collection. UK Biobank has full ethical approval from the NHS National Research Ethics Service (16/NW/0274).

## Figure legends

Checklist S1. Strengthening the Reporting of Observational Studies in Epidemiology (STROBE)

Figure S1. Flow chart of participants included in main analysis

Figure S2. Association between employment status and odds of severe COVID-19, stratified by a) sex and b) ethnicity, additionally controlled for self-reported sleep duration.

Model (a) adjusted for: age, sex (for all participants), Townsend score, BMI, cancer (self-reported, past or current at time of data collection), co-morbidities (yes/no), smoking status (never, previous current), ethnicity, and self-reported sleep duration. Model (b) same as model (a) except without ethnicity.

Table S1. Unadjusted Associations between employment status and odds of severe COVID-19

