## Supplementary for "Association of working shifts, inside and outside of healthcare, with risk of severe COVID-19: An observational study"

### Checklist S1. Strengthening the Reporting of Observational Studies in Epidemiology (STROBE)

| | Item No. | Recommendation | Page No. |
|---|---|---|---|
| **Title and abstract** | 1 | (*a*) Indicate the study's design with a commonly used term in the title or the abstract | Title |
| | | (*b*) Provide in the abstract an informative and balanced summary of what was done and what was found | Abstract |
| **Introduction** | | | |
| Background/rationale | 2 | Introduction | Introduction, paragraphs 1-3 |
| Objectives | 3 | State specific objectives, including any prespecified hypotheses | Introduction, paragraph 3 |
| **Methods** | | | |
| Study design | 4 | Present key elements of study design early in the paper | Methods, study population |
| Setting | 5 | Describe the setting, locations, and relevant dates, including periods of recruitment, exposure, follow-up, and data collection | Methods, study population |
| Participants | 6 | (*a*) *Cohort study*—Give the eligibility criteria, and the sources and methods of selection of participants. Describe methods of follow-up | Methods, study population, exposure, outcome |
| | | (*b*) *Cohort study*—For matched studies, give matching criteria and number of exposed and unexposed | N/A |
| Variables | 7 | Clearly define all outcomes, exposures, predictors, potential confounders, and effect modifiers. Give diagnostic criteria, if applicable | Methods, exposure, outcome |
| Data sources/ measurement | 8* | For each variable of interest, give sources of data and details of methods of assessment (measurement). Describe comparability of assessment methods if there is more than one group | Methods, exposure, outcome, co-variates/confounders |
| Bias | 9 | Describe any efforts to address potential sources of bias | Statistical analysis, sensitivity analyses |
| Study size | 10 | Explain how the study size was arrived at | Supplementary information, Figure S1 |

Continued on next page

| | | | |
|---|---|---|---|
| Quantitative variables | 11 | Explain how quantitative variables were handled in the analyses. If applicable, describe which groupings were chosen and why | Methods, statistical analysis |
| Statistical methods | 12 | (*a*) Describe all statistical methods, including those used to control for confounding | Methods, statistical analysis |
| | | (*b*) Describe any methods used to examine subgroups and interactions | Methods, statistical analysis, sensitivity analyses |
| | | (*c*) Explain how missing data were addressed | Methods, statistical analysis |
| | | (*d*) *Cohort study*—If applicable, explain how loss to follow-up was addressed | N/A |
| | | (*e*) Describe any sensitivity analyses | Methods, statistical analysis, sensitivity analyses |
| **Results** | | | |
| Participants | 13* | (a) Report numbers of individuals at each stage of study—eg numbers potentially eligible, examined for eligibility, confirmed eligible, included in the study, completing follow-up, and analysed | Methods, study population |
| | | (b) Give reasons for non-participation at each stage | Methods, study population |
| | | (c) Consider use of a flow diagram | Supplementary information, Figure S1 |
| Descriptive data | 14* | (a) Give characteristics of study participants (eg demographic, clinical, social) and information on exposures and potential confounders | Results paragraph 1, Table 1 |
| | | (b) Indicate number of participants with missing data for each variable of interest | Supplementary information, Figure S1 |
| | | (c) *Cohort study*—Summarise follow-up time (eg, average and total amount) | N/A |
| Outcome data | 15* | *Cohort study*—Report numbers of outcome events or summary measures over time | Results, paragraphs 2-3, Table 1 |
| Main results | 16 | (*a*) Give unadjusted estimates and, if applicable, confounder-adjusted estimates and their precision (eg, 95% confidence interval). Make clear which confounders were adjusted for and why they were included | Results paragraphs 3-5, Figures 1, 2, Supplementary information Table S1 and Figure S2, |
| | | (*b*) Report category boundaries when continuous variables were categorized | N/A |
| | | (*c*) If relevant, consider translating estimates of relative risk into absolute risk for a meaningful time period | N/A |

Continued on next page

| | | | |
|---|---|---|---|
| Other analyses | 17 | Report other analyses done—eg analyses of subgroups and interactions, and sensitivity analyses | Results, sensitivity analyses Figure 2, Supplementary information Figure S2 |
| **Discussion** | | | |
| Key results | 18 | Summarise key results with reference to study objectives | Discussion paragraph 1 |
| Limitations | 19 | Discuss limitations of the study, taking into account sources of potential bias or imprecision. Discuss both direction and magnitude of any potential bias | Discussion paragraph 5 |
| Interpretation | 20 | Give a cautious overall interpretation of results considering objectives, limitations, multiplicity of analyses, results from similar studies, and other relevant evidence | Discussion paragraphs 5-6 |
| Generalisability | 21 | Discuss the generalisability (external validity) of the study results | Discussion, paragraph 5 |
| **Other information** | | | |
| Funding | 22 | Give the source of funding and the role of the funders for the present study and, if applicable, for the original study on which the present article is based | End of manuscript |

*Give information separately for cases and controls in case-control studies and, if applicable, for exposed and unexposed groups in cohort and cross-sectional studies.

**Note:** An Explanation and Elaboration article discusses each checklist item and gives methodological background and published examples of transparent reporting. The STROBE checklist is best used in conjunction with this article (freely available on the Web sites of PLoS Medicine at http://www.plosmedicine.org/, Annals of Internal Medicine at http://www.annals.org/, and Epidemiology at http://www.epidem.com/). Information on the STROBE Initiative is a

**Figure S1: Flow chart of participants included in the study**

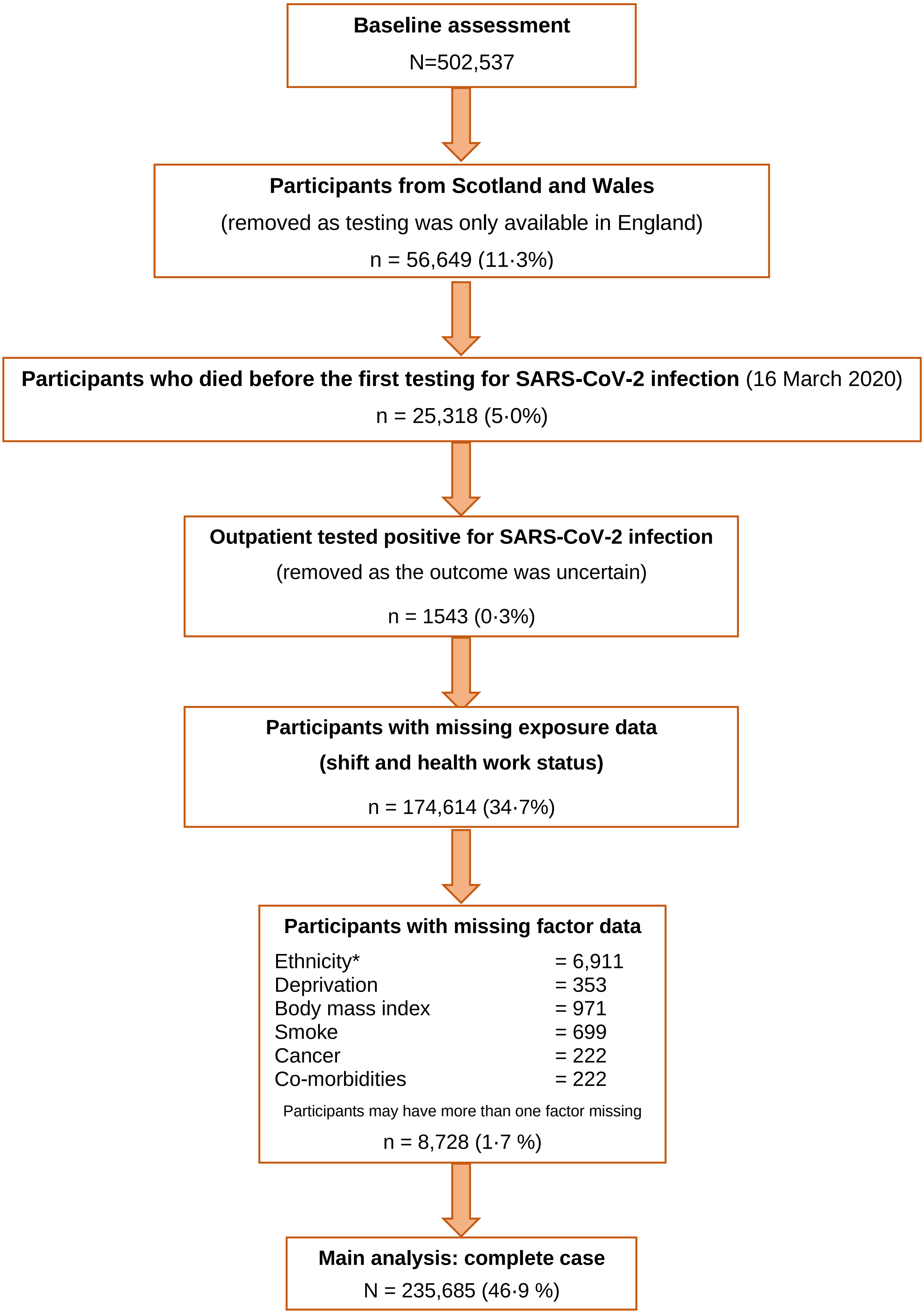

* White Ethnicity, South Asian, Black and African Caribbean

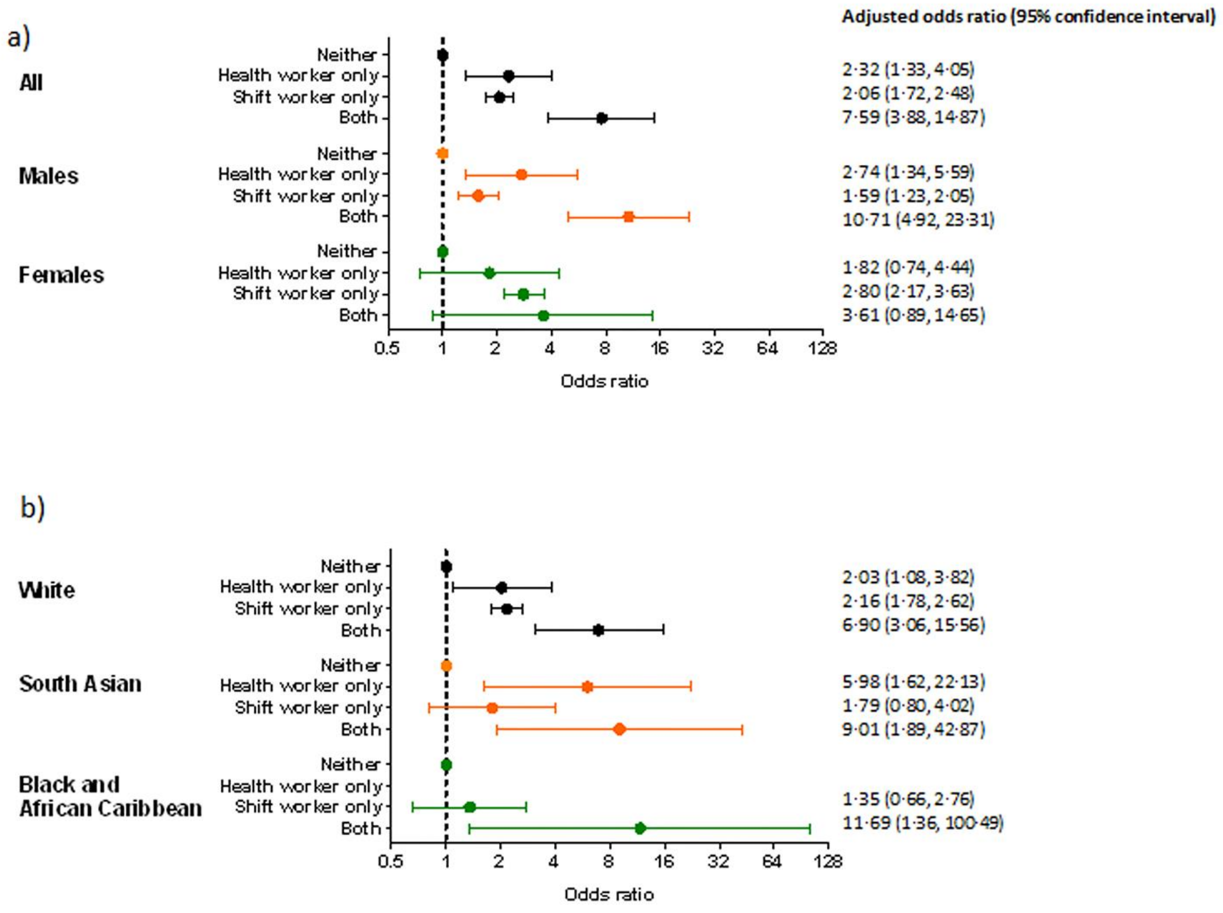

Figure S2. Association between employment status and odds of severe COVID-19, stratified by a) sex and b) ethnicity, additionally controlled for self-reported sleep duration.

**Table S1. Unadjusted Associations between employment status and odds of severe COVID-19 (Odds ratio (95% confidence interval))**

| **Main analysis** | **All** | **Men** | **Women** |
|---|---|---|---|
| ***Severe covid-19*** | | | |
| Neither | Reference | Reference | Reference |
| Health worker only | 1·99 (1·14, 3·46) | 2·43 (1·20, 4·93) | 1·56 (0·64, 3·81) |
| Shift worker only | 2·45 (2·06, 2·90) | 1·85 (1·45, 2·36) | 3·26 (2·56, 4·15) |
| Both | 7·79 (4·14, 14·66) | 11·56 (5·70, 23·81) | 3·34 (0·83, 13·53) |
| | **White European** | **South Asian** | **Black and African Caribbean** |
| ***Severe covid-19*** | | | |
| Neither | Reference | Reference | Reference |
| Health worker only | 1·80 (0·96, 3·38) | 4·60 (1·31, 16·19) | - |
| Shift worker only | 2·41 (1·99, 2·90) | 1·96 (0·89, 4·32) | 1·60 (0·79, 3·24) |
| Both | 6·13 (2·72, 13·80) | 7·14 (1·59, 32·10) | |
| **Under 66 y** | **All** | | |
| ***Severe covid-19*** | | | |
| Neither | Reference | | |
| Health worker only | 3·24 (1·71, 6·14) | | |
| Shift worker only | 3·12 (2·49, 3·91) | | |
| Both | 13·17 (6·92, 25·06) | | |
| **Over 66 y** | **All** | | |
| ***Severe covid-19*** | | | |
| Neither | Reference | | |
| Health worker only | 0·88 (0·28, 2·74) | | |
| Shift worker only | 1·78 (1·35, 2·35) | | |
| Both | - | | |
